# Feasibility study of a novel, low-cost splint device for children with foot drop

**DOI:** 10.64898/2026.05.07.26352389

**Authors:** Timothy A. Exell, Joseph M. Moore, Amy Wright, Susan Cleverley, Joshua Roel Ferreira, Rebecca Williams, Zoe L. Saynor

## Abstract

**Importance:** Foot drop impairs mobility for many children globally, causing life-long health issues. Existing treatments are costly, custom-made, and require frequent clinical visits. A new, low-cost, off-the-shelf splint (OrthoPed) could improve access and user experience.

**Objective:** To determine the feasibility of recruiting children (4–17 years) with moderate foot drop and collecting biomechanical, clinical, and patient-reported outcomes to compare OrthoPed with existing treatments.

**Design:** Single-centre cross-sectional feasibility and pilot study informing a future randomised clinical trial.

**Participants:** Twelve children (target=20; mean age=10.6 ± 3.5 years; 2 females) with moderate foot drop and prescribed orthotic support were recruited via physiotherapy.

**Intervention:** The new OrthoPed splint was compared against existing treatments: ankle foot orthoses (AFOs) and Lycra socks.

**Main outcome measures:** Primary outcome: recruitment and retention rates. Secondary outcomes: biomechanical and clinical gait measures, alongside useability and performance questionnaires.

**Results:** Recruitment reached 22% of eligible participants (an “amber” rating for future trials). Despite four dropouts due to treatment burden, all outcome measures were successfully collected. Preliminarily, OrthoPed supported more natural gait mechanics than AFOs and offered better usability and comfort than AFOs and Lycra socks, potentially enhancing adherence.

**Conclusions:** Recruiting children for orthotic trials is feasible, though coordinating gait testing with routine clinical appointments could improve future recruitment. Importantly, low-cost orthotic devices may provide better usability, accessibility and adherence than existing prescribed options.

**Clinical Trial Registration number:** NCT05587010

**Contribution of the Paper:**

- The OrthoPed splint is a safe, feasible, low-cost alternative for paediatric foot drop, balancing gait support with natural ankle mobility.
- A definitive study will addresses a critical gap in neurorehabilitation research for children.
- Findings suggest OrthoPed supports more natural mechanics and preserved movement compared to motion-restricting rigid AFOs.
- The off-the-shelf design improves patient experience through superior comfort and footwear compatibility, likely enhancing treatment adherence.
- By eliminating bespoke fitting and frequent clinic visits, OrthoPed can reduce healthcare costs and clinical workloads, offering a transformative global solution for foot drop management.

## INTRODUCTION

Foot drop impairs gait and mobility in children, often leading to reduced independence and quality of life ^1^ and long-term development. Ankle-foot orthoses (AFOs) are commonly prescribed to manage foot drop by supporting the ankle during gait ^2,3^. However, conventional AFOs have well-recognised limitations: while they reduce plantarflexion during the gait swing phase, they can suppress dorsiflexor muscle activity ^3–6^, and restrict some movement patterns, limiting tasks such as stair climbing and floor transitions ^7^. Importantly, discomfort often leads to poor adherence to AFOs ^8^.

Lycra socks offer an alternative for moderate foot drop in children and adolescents ^9^, allowing for greater ankle movement than AFOs but with inconsistent gait improvements and limited structural support ^10^. Additionally, both AFOs and Lycra socks typically require bespoke fitting and multiple clinic visits, adding strain on health services and families. These devices can also represent substantial costs, with estimates exceeding £1,800 per child annually.

OrthoPed is a novel, low-cost, off-the-shelf elastic splint designed to prevent plantarflexion while preserving natural ankle movement. Its lightweight, comfortable design allows direct provision without the need for custom fitting or repeated appointments. This device aims to offer substantial time and cost efficiencies, with potentially improved patient experience and compliance.

Despite the clear clinical need and promising rationale, no formal studies have evaluated the feasibility of using such a device in children and adolescents with foot drop. Paediatric rehabilitation trials pose unique challenges ^11^, particularly in recruitment, retention, and collecting biomechanical outcomes in young, often medically complex participants. Quantifying movement-related outcomes in this group also demands careful methodological planning, given the burden of high-tech gait laboratory assessments.

This feasibility study investigated the recruitment, retention, and safety of children with foot drop comparing the OrthoPed splint against three conditions: barefoot, AFO, and Lycra sock. By gathering preliminary gait data, these findings inform a future definitive trial evaluating the clinical efficacy, acceptability, and cost-effectiveness of this novel paediatric intervention.

## METHODS

### Ethical Approval

This study was pre-registered on a clinical trials database (clinicaltrials.gov, study number NCT05587010). Ethical approval was provided by Yorkshire and Humber – Sheffield NHS Research Ethics Committee (reference 23/YH/0004) and the Health Research Authority along with confirmation of no-objection from the Medicines and Healthcare products Regulatory Agency. All participants aged < 16 years provided assent and had written informed consent provided by a caregiver, participants aged 16-17 years provided written informed consent.

### Study Design and Participants

This single-centre cross-sectional pilot study recruited children aged 4–17 with moderate foot drop, defined as a minimum of 5 degrees of passive dorsiflexion, were eligible. This level of ankle mobility was required for use of the new OrthoPed splint. Patients were identified through their physiotherapy care. Participants attended three visits: initial measurement, university-based biomechanical gait analysis, and clinic-based clinical measures and feedback.

Exclusion criteria included: visual impairment when corrected to not be able to see targets when walking, unable to understand or cooperate with study protocol, health contraindications to exercise e.g. cardiac disease, significant equinovarus / valgus, hypertonicity, weakness deformity, dominant toe walkers.

### Intervention and Comparators

The intervention was a new low-cost, off-the-shelf splint device, the “OrthoPed splint” worn with participants’ own footwear. Comparators included existing common treatments for foot drop of individualised rigid ankle foot orthoses (AFO) and Lycra socks, worn with participants own footwear. A barefoot condition was also included as a baseline. The three support devices used in the study are shown in Figure 1.

**Figure 1:**
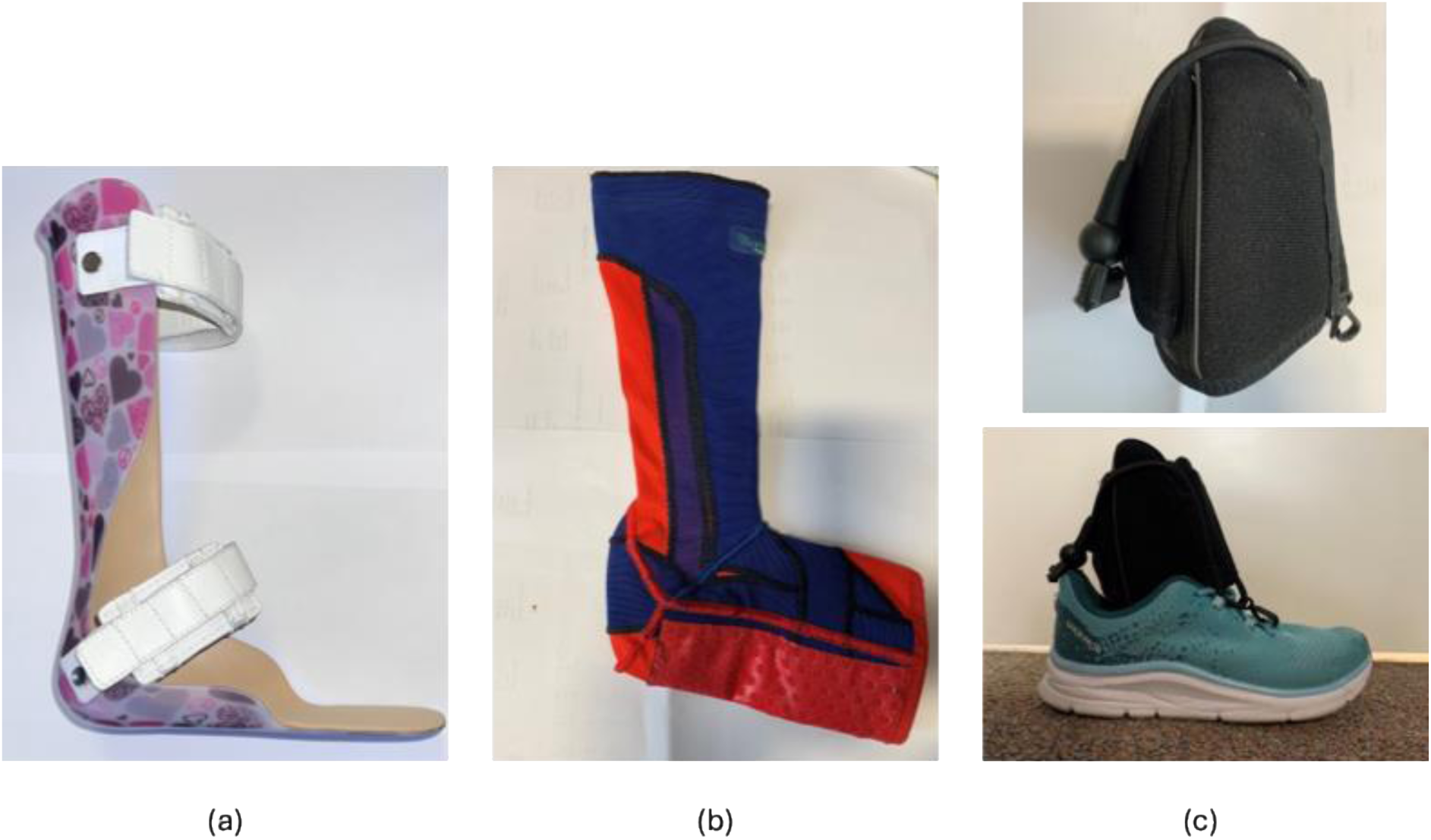
Existing orthotic devices: AFO (a) and Lycra sock (b); the new OrthoPed splint (c).

### Data Collection

#### Measurement and Fitting of Orthotic Supports

Participants attended an initial visit (Visit 1) at the Children’s physiotherapy gym where they were measured for the AFO, Lycra sock and OrthoPed orthotics. Measurements were taken by a trained orthotist or physiotherapist with experience of measuring for these devices.

#### Biomechanical Data Collection

On Visit 2, participants attended the Movement Analysis and Rehabilitation Lab at the University of **\*\*\***. Participants’ mass and height were measured using a stadiometer (Seca Ltd, Leicester, UK) and weighing scales (Seca Ltd, Leicester, UK). Fifty reflective markers were then attached to their skin or clothing using adhesive tape by experienced clinical gait biomechanists. Markers were attached bilaterally to 1st, 3rd and 5th metatarsal-phalangeal joints, calcaneus, medial and lateral ankle malleoli, medial and lateral femoral condyles, lateral to the greater trochanter, superficial to the anterior superior iliac spine, iliac crest and posterior superior iliac spine, superior to the acromioclavicular joint, medial and lateral humeral condyles, radial and ulnar styloid processes, four markers on the anterior shank and anterior thigh.

Biomechanical data were collected using an optoelectronic motion capture system (Qualisys, 250 Hz) and Kistler force plates (1000 Hz). A lower-limb model quantified joint angles, step characteristics, and ground reaction forces across randomised support conditions. Participants completed five walking trials along a 6 m walkway at a self-selected speed, without intentionally targeting the centrally located force plates. Clinical measures included the six-minute walk test (6MWT) ^12^, single-leg stance ^13^, Timed Up and Go (TUG) ^14^, and Gait Assessment and Intervention Tool (GAIT) ^15^, and device donning time.

Following clinical measures, participants and parents completed the Peds-QL^TM^ Measurement Model for the Pediatric Quality of Life Inventory^TM^ (Peds-QL) questionnaire ^16^ and a questionnaire on device fit, comfort, and performance. A separate parent questionnaire was distributed after data collection, seeking feedback on the new device.

### Data Analysis

#### Biomechanical Data

A pelvis and lower-limb model in Visual 3D (HAS Motion) allowed calculation of joint angles as relative rotations between adjacent segments. Joint angle data were exported throughout the gait cycle. Touchdown instants were identified using a vertical force threshold or vertical acceleration of the calcaneus marker. Step length and frequency were determined for each step.

### Outcome Measures

#### Feasibility

Feasibility was assessed against red/amber/green progression criteria ^17^ focusing on recruitment and retention rates (Table 2). Acceptability was evaluated through participant and parent questionnaires regarding comfort, usability, and device preference. Progression criteria for a definitive trial were informed by local data on annual eligible patients, setting minimum recruitment thresholds for future clinical trials. This study’s progression criteria were defined as:

- Red: stop; intractable issues.
- Amber: amend; remediable issues, proceed with caution (e.g., adapt testing for redesign)
- Green: continue; no concerning issues threaten future trial success.

These pre-specified criteria judged future clinical trial viability within planned timetable and budget, addressing key uncertainties or risks ^17^.

#### Data Collection Feasibility and Completeness

Completion rates for all planned assessments. Data quality and completeness (e.g., missing data points, any issues with data collection), practicality of using motion capture cameras and force plate with children and young people recruited to the study, number of trials performed to achieve the required successful trials for each condition.

#### Safety and Tolerability

Incidence and type of adverse events or discomfort related to the OrthoPed splint.

Reasons for early discontinuation due to adverse events.

#### Preliminary Clinical and Biomechanical Outcomes (to inform future sample size)

Outcome measures collected during each support condition were defined as:

- Six-minute walk test: The total walking distance covered during 6 minutes.
- The Single Leg Stance (SLS) Test: Total time from the non-balancing foot leaving the ground until the foot touches the ground again or the arms leave the hips.
- Timed Up and Go: The time taken from the participant to stand from a chair (time started when knees straight) to walk to a mark in the floor 3 m away, turn and walk back to the seat and return to sitting (time ends).
- Gait Assessment and Intervention Tool: The gait score given by the clinician using the Gait Assessment and Intervention Tool criteria
- Time taken to put on each device 1 handed: The time taken to complete this was recorded as well as whether they were able to put the device on independently or if they required assistance.
- Sagittal plane joint angles: ankle, knee and hip joints at the times of touchdown, as well as minimum, maximum and range of motion for each joint throughout the gait cycle.
- Ground Reaction Force: peak vertical ground reaction force during stance for each side of the body.

#### Patient/ parent reported outcome measures

For the device questionnaire, response values were summed for Questions 1-11 about the performance of each orthotic device to give an overall score. Scores were separately summed for Questions 12 – 15 for the devices that were not the participants’ usual orthotic devices. PedsQL data were scored according to the Scaling and Scoring of the Pediatric Quality of Life Inventory guidelines. Where responses were reverse scored (0 = 100% through to 4 = 0%) and then mean scored within each response category.

#### Statistical analyses

As a feasibility pilot trial, analyses were primarily descriptive. For device questionnaire data, ordinal data were analysed using Friedman’s ANOVA to compare for differences between support conditions. For conditions showing a significant effect (p<0.05), post-hoc pairwise Friedman’s comparisons were conducted.

## RESULTS

### Feasibility

Recruitment took place October 2023 - March 2024 and was stopped due to the funding award end date. Participant demographics are displayed in Table 1. Twelve children initially consented; eight completed the trial. Recruitment was 22% (“Amber”), largely due to the travel burden for medically complex oncology participants. However, retention was 100% (“Green”), and no adverse events occurred. All planned clinical and biomechanical data were successfully collected for all but the youngest participant (5-9 Years old), for whom fewer gait trials were collected due to difficulty following the protocol instructions.

**Table 1:**
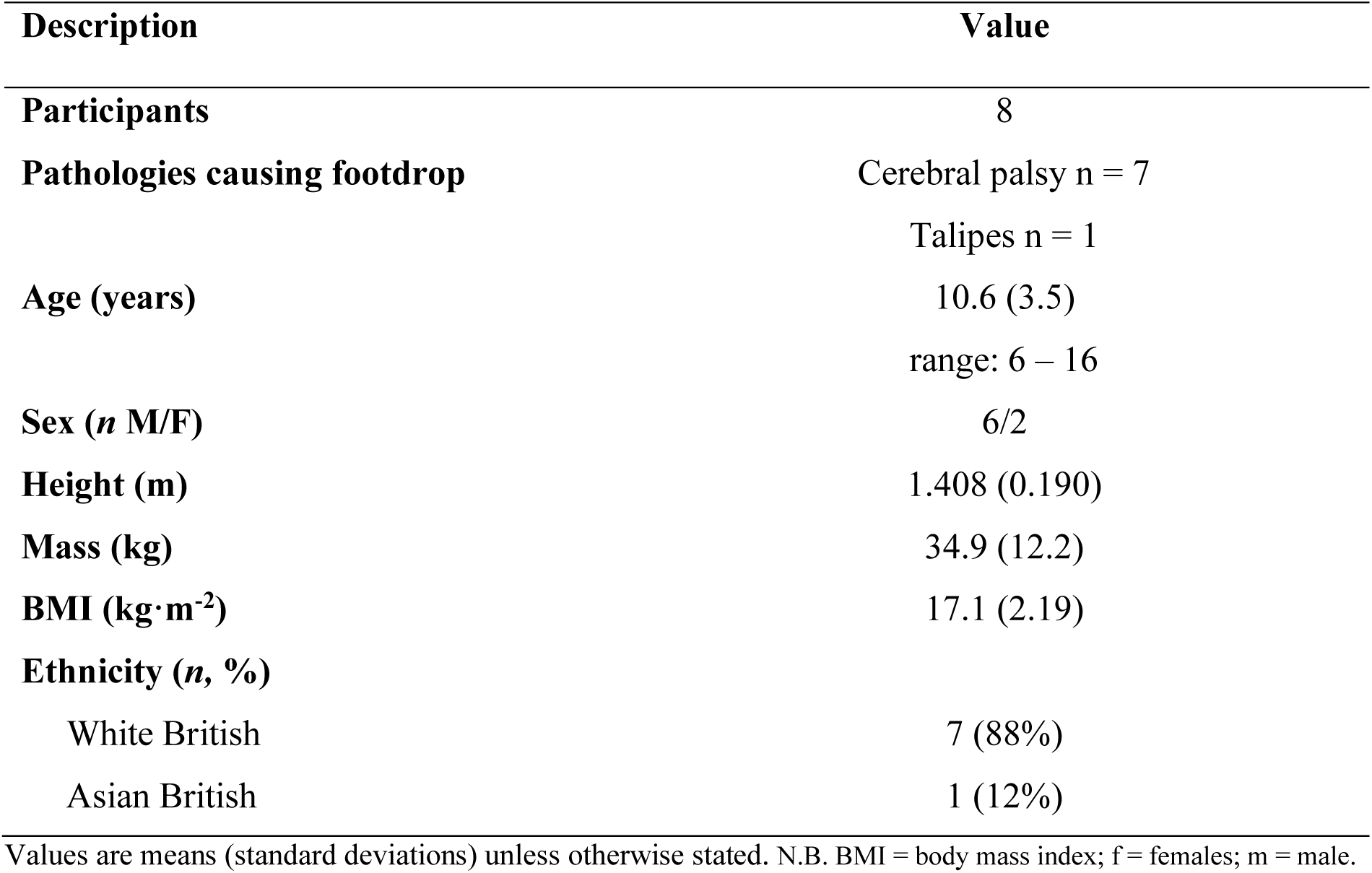
Participant demographics.

**Table 2.**
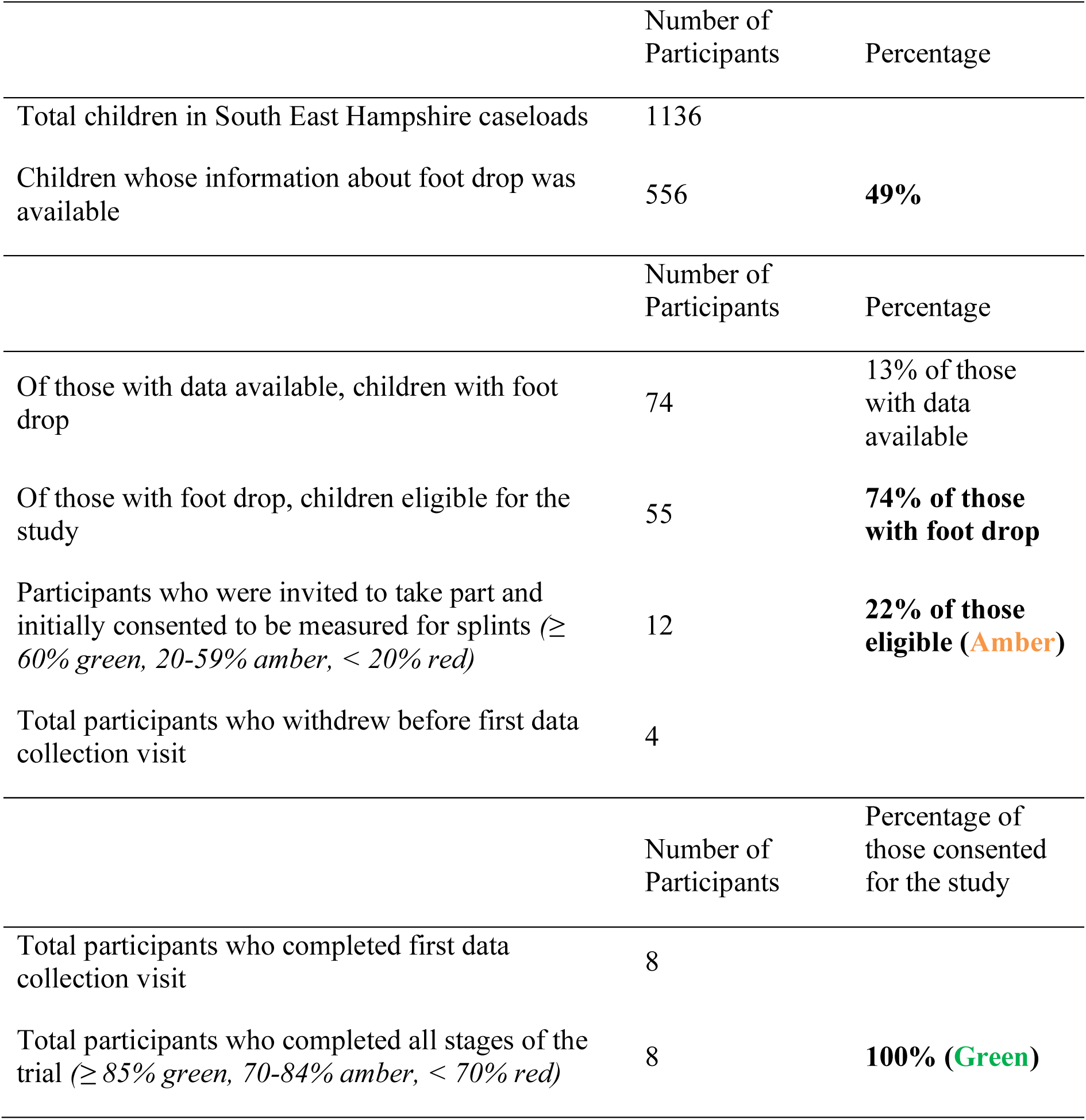
Feasibility results of recruiting and retaining participants to the study against pre-defined trial progression criteria.

|  | Number of<br>Participants | Percentage |
| --- | --- | --- |
| Total children in South East Hampshire caseloads | 1136 |  |
| Children whose information about foot drop was available | 556 | **49%** |
|  | Number of<br>Participants | Percentage |
| Of those with data available, children with foot drop | 74 | 13% of those with data available |
| Of those with foot drop, children eligible for the study | 55 | **74% of those with foot drop** |
| Participants who were invited to take part and initially consented to be measured for splints ($\geq 60\%$ green, 20-59% amber, $< 20\%$ red) | 12 | **22% of those eligible (Amber)** |
| Total participants who withdrew before first data collection visit | 4 |  |
|  | Number of<br>Participants | Percentage of<br>those consented<br>for the study |
| Total participants who completed first data collection visit | 8 |  |
| Total participants who completed all stages of the trial ($\geq 85\%$ green, 70-84% amber, $< 70\%$ red) | 8 | **100% (Green)** |

### Acceptability and Useability

Device questionnaire responses (median and inter-quartile range) are in Table 3. OrthoPed and Lycra socks scored significantly higher than AFOs for comfort and shoe compatibility. Parents rated OrthoPed highly for its discrete design and perceived walking benefits, with 5/6 reporting their child was “likely” to wear it frequently. While one-handed donning was difficult across all devices, OrthoPed was rated easier to use than the Lycra sock.

**Table 3.** Median (IQR) responses to the device questionnaire, answered for each orthotic support condition. Responses available for each question were 1 = Completely disagree, 2 = Somewhat disagree, 3 = Neutral, 4 = Somewhat agree, 5 = Completely agree. Shaded cells indicate a significant difference with superscript letters showing which conditions the value is significantly different to.

|  | AFO | Lycra sock | OrthoPed |
| --- | --- | --- | --- |
| 1. Overall, the device is easy to use | 4.00 (0.75) | 3.00 (1.75) <sup>O</sup> | 5.00 (1.75) <sup>L</sup> |
| 2. Overall, the device is simple to use | 4.00 (0.75) | 3.00 (2.00) | 4.50 (2.75) |
| 3. The device is easy to put on with one hand | 1.00 (2.50) | 1.00 (0.00) | 1.00 (0.75) |
| 4. The device can be easily worn with shoes | 3.00 (1.50) <sup>LO</sup> | 5.00 (0.00) <sup>A</sup> | 5.00 (0.00) <sup>A</sup> |
| 5. The device is comfortable in general | 2.50 (2.75) <sup>LO</sup> | 5.00 (2.00) <sup>A</sup> | 5.00 (1.00) <sup>A</sup> |
| 6. The device is comfortable when walking | 1.50 (3.25) <sup>LO</sup> | 5.00 (1.75) <sup>A</sup> | 4.50 (1.00) <sup>A</sup> |
| 7. It is easy to remove the device | 5.00 (0.00) <sup>LO</sup> | 2.00 (1.50) <sup>AO</sup> | 3.50 (1.00) <sup>AL</sup> |
| 8. I feel confident in my ability to use the device | 4.00 (1.75) | 4.00 (0.75) | 5.00 (2.75) |
| 9. It is easier to walk when wearing the device | 2.00 (3.50) <sup>L</sup> | 4.00 (1.50) <sup>A</sup> | 4.50 (1.75) |
| 10. I can walk faster when wearing the device | 1.50 (3.75) <sup>LO</sup> | 4.00 (2.75) <sup>A</sup> | 5.00 (1.50) <sup>A</sup> |
| 11. I feel more stable walking when wearing the device | 3.00 (3.50) | 3.50 (2.75) | 4.50 (2.75) |
| 12. I find the device easier to put on than my usual orthotic support | 3.00 (4.00) | 1.00 (0.50) | 2.00 (3.50) |
| 13. I find the device more comfortable than my usual orthotic support | 3.00 (3.50) | 5.00 (3.00) | 5.00 (2.50) |
| 14. I prefer the support of the device compared with my usual orthotic support | 3.00 (4.00) | 3.00 (3.50) | 5.00 (2.50) |
| 15. I prefer the fit of the device compared with my usual orthotic support | 2.00 (4.00) | 3.00 (3.50) | 5.00 (2.50) |

Feedback on the OrthoPed splint from six parents is included in Table 4. Most participants were ‘quite likely’ or ‘very likely’ to wear the splint frequently. Identified practical benefits include wider shoe compatibility and a discrete design allowing other clothing to be worn. Only two responses indicated that the user was able to put the device on themselves one-handed, yet most parents found it ‘very easy’ or ‘quite easy’ to put on and take off the splint. Five parents perceived a walking benefit when using the OrthoPed splint compared to walking barefoot.

**Table 4.**
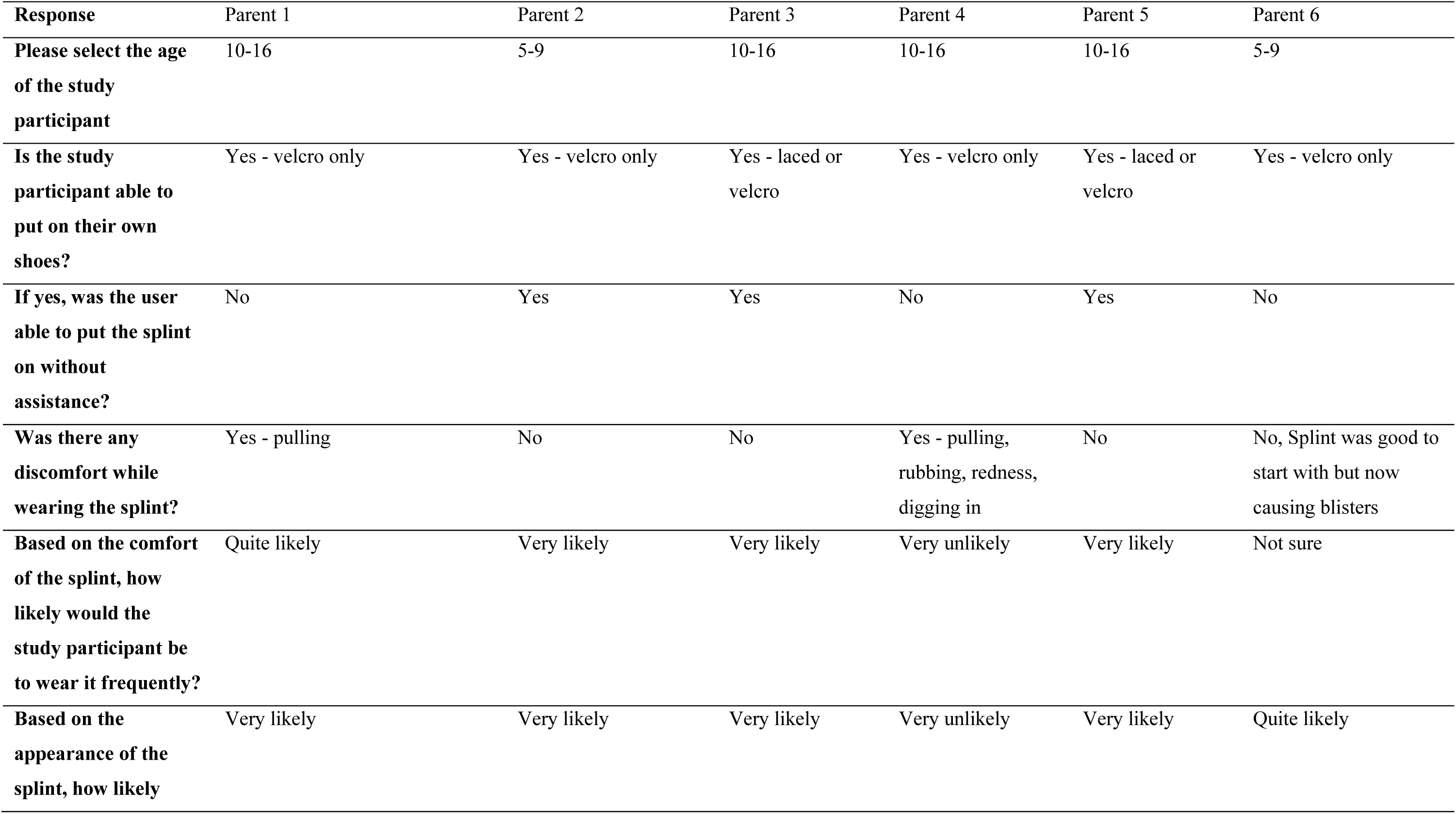
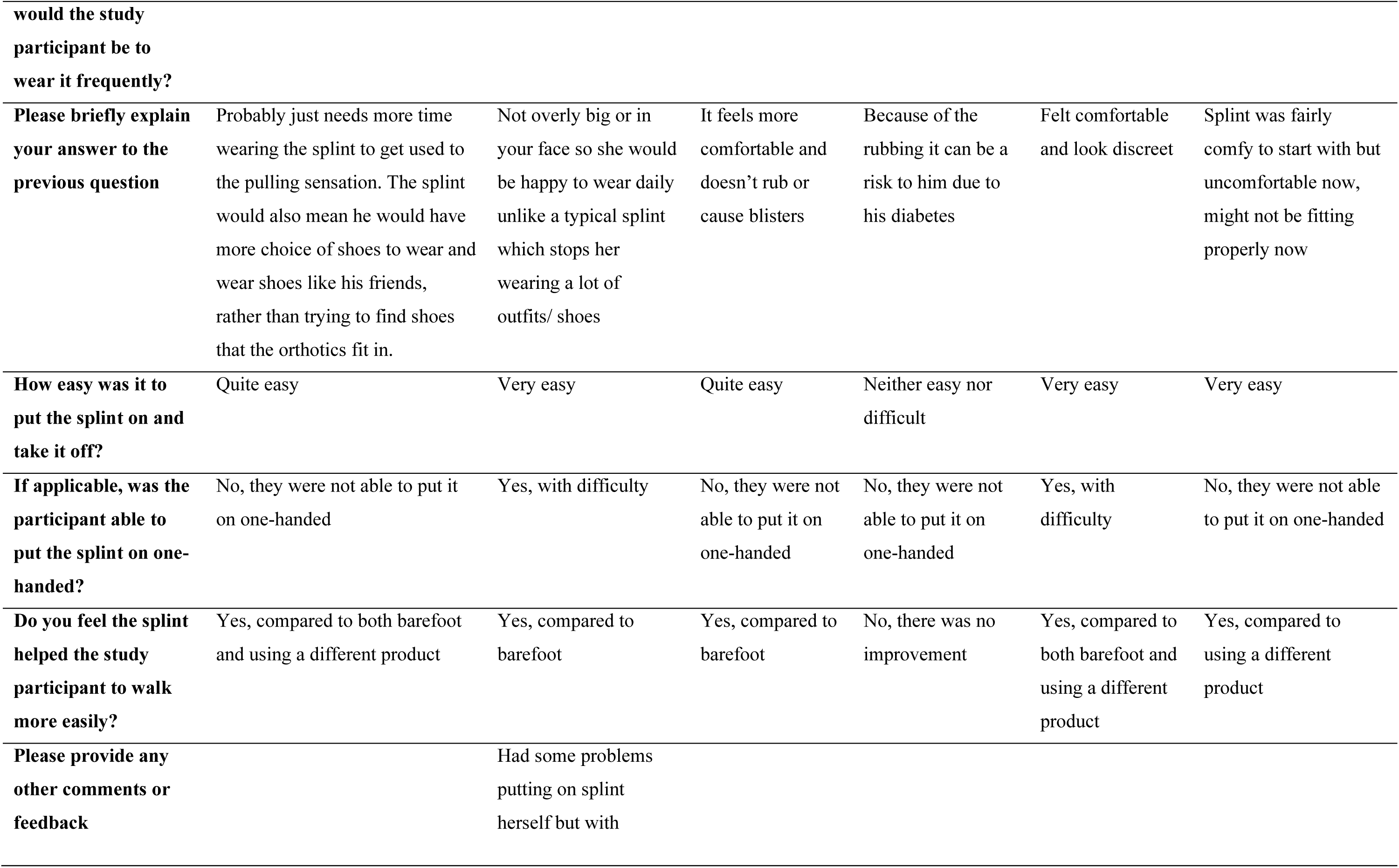
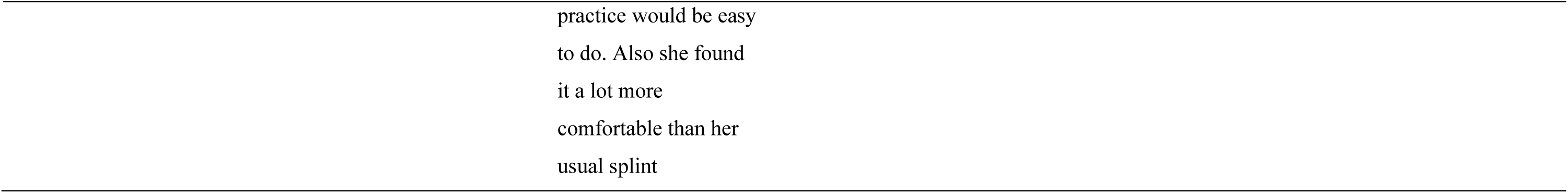
Parent feedback on use of the OrthoPed splint.

### Preliminary Clinical Outcome Measures

Group outcomes for clinical gait tests are shown in Table 5. OrthoPed yielded the highest mean distance in the 6MWT, while AFOs performed worst in both 6MWT and TUG. AFOs provided the highest stability in balance tests. However, variability between participants within each condition was large compared to the group mean differences across all tests. No harms were detected in clinical outcome measures

**Table 5.** Mean (SD) clinical gait measures.

|  | **Barefoot** | **AFO** | **Lycra sock** | **OrthoPed** |
| --- | --- | --- | --- | --- |
| **Six-minute walk (m)** | 363 (70) | 7.2 (1.5) | 7.2 (1.5) | 20.6 (11.2) |
| **Single leg balance (s)** | 309 (123) | 11.6 (6.8) | 11.6 (6.7) | 15.4 (7.9) |
| **Timed up and go (s)** | 332 (162) | 7.0 (2.0) | 7.0 (2.0) | 19.4 (12.6) |
| **GAIT (lowest score favoured)** | 401 (71) | 7.1 (2.2) | 7.1 (2.2) | 17.4 (12.0) |

Note: AFO = ankle foot orthosis; GAIT = Gait Assessment and Intervention Tool.

### Preliminary Biomechanical Outcome Measures

Tables 6 and 7 contain step characteristics and ground reaction force results and joint angle data for affected sides, respectively. OrthoPed and Lycra socks allowed significantly more ankle range of motion and plantarflexion than the AFO, which restricted movement most severely. Ground reaction force data, were similar across all support conditions, within 4% and 8% bodyweight for affected and non-affected limbs, respectively. Peak force values were more consistent for OrthoPed and Lycra sock conditions (lower SD) than for barefoot and AFO.

**Table 6.** Mean (SD) step length (SL), step frequency (SF) and step velocity (SV) for steps from affected foot to non-affected foot ground contact (A-NA) and steps from non-affected foot to affected foot ground contact (NA-A) and peak vertical ground reaction force (vGRF) for ground contact phases of the affected (A) and non-affected (NA) sides.

| Test condition |  | Barefoot | AFO | Lycra sock | OrthoPed |
| --- | --- | --- | --- | --- | --- |
| SL (m) | A-NA | 0.54<br>(0.08) | 0.54<br>(0.19) | 0.50<br>(0.03) | 0.52<br>(0.18) |
|  | NA-A | 0.50<br>(0.04) | 0.55<br>(0.13) | 0.53<br>(0.09) | 0.57<br>(0.14) |
| SF (Hz) | A-NA | 124<br>(16) | 120<br>(17) | 128<br>(16) | 125<br>(17) |
|  | NA-A | 118<br>(15) | 105<br>(23) | 114<br>(12) | 101<br>(18) |
| SV (m/s) | All steps | 1.03<br>(0.16) | 1.01<br>(0.34) | 1.11<br>(0.21) | 1.10<br>(0.24) |
| vGRF<br>(BW) | A | 138<br>(26) | 138<br>(36) | 139<br>(22) | 135<br>(17) |
|  | NA | 136<br>(26) | 132<br>(30) | 134<br>(17) | 140<br>(20) |

**Table 7.** Mean (SD) sagittal plane joint angle data for the ankle, knee, hip and elbow joints of the affected side. Shaded cells indicate a difference between conditions at least as large as the standard deviation for those conditions, with superscript letters showing which conditions the value is different to. Larger positive values indicate greater hip and knee flexion and ankle dorsiflexion (0° = full extension).

| Measure | Joint | Barefoot | AFO | Lycra sock | OrthoPed |
| --- | --- | --- | --- | --- | --- |
| Touchdown<br>angle<br>(°) | Ankle | 61<br>(9) | 70<sup>LO</sup><br>(6) | 60<sup>A</sup><br>(6) | 64<sup>A</sup><br>(5) |
|  | Knee | 25<br>(8) | 25<br>(8) | 20<br>(8) | 21<br>(8) |
|  | Hip | 41<br>(9) | 44<br>(7) | 39<br>(9) | 41<br>(6) |
| Peak flexion<br>(°) | Ankle | 48<br>(13) | 64<sup>LO</sup><br>(6) | 42<sup>A</sup><br>(8) | 53<sup>A</sup><br>(7) |
|  | Knee | 77<br>(8) | 76<br>(14) | 80<br>(10) | 78<br>(12) |
|  | Hip | -5<br>(8) | -2<br>(12) | -4<br>(7) | -5<br>(8) |
|  | Elbow | 120 | 113 | 126 | 116 |

|  |  | (23) | (35) | (20) | (35) |
| --- | --- | --- | --- | --- | --- |
| Peak<br>extension<br>(°) | Ankle | 85<br>(9) | 83<br>(6) | 85<br>(8) | 87<br>(7) |
|  | Knee | 4<br>(6) | 6<br>(9) | 6<br>(9) | 3<br>(7) |
|  | Hip | 52<br>(11) | 51<br>(8) | 52<br>(7) | 52<br>(10) |
|  | Elbow | 154<br>(18) | 146<br>(29) | 157<br>(14) | 156<br>(19) |
| Range of<br>motion<br>(°) | Ankle | 37<sup>A</sup><br>(12) | 19<sup>BLO</sup><br>(5) | 41<sup>AO</sup><br>(13) | 33<sup>AL</sup><br>(11) |
|  | Knee | 72<br>(12) | 70<br>(20) | 71<br>(18) | 73<br>(22) |
|  | Hip | 57<br>(10) | 53<br>(13) | 54<br>(11) | 56<br>(15) |

PedsQL questionnaire responses from children with foot drop and their parents, revealing the condition’s wide-ranging impact. Physical functioning showed the highest impact, with all median scores for orthotic users being <54%. Individual scores also indicated significant impact on emotional, social, and school functioning. Family impact was generally lower, but some scores highlighted an influence on physical and emotional functioning, communication, worry, daily activities, and family relationships due to living with someone with foot drop.

## DISCUSSION

This is the first study to evaluate the feasibility and acceptability of OrthoPed, a novel, market-approved, low-cost paediatric foot drop device. Our results demonstrate strong support for the device among children, parents, and clinicians, highlighting its comfort, usability, and promising impact on gait outcomes.

The study showed 100% retention and high acceptability of procedures. Although recruitment fell below target (22%) due to the travel burden on medically complex oncology and neurology patients, these insights are critical for future trial design. Embedding outcome collection within routine clinical care, such as oncology or orthotics clinics, would likely enhance accessibility and participation.

OrthoPed was well-tolerated with no adverse events. While biomechanical data were successfully collected across all conditions, concentration difficulties in younger participants suggest that future trials should reduce the number of repetitions per condition to minimise fatigue.

Functionally, OrthoPed demonstrated superior performance in the 6MWT and strong GAIT scores while maintaining ankle movement closer to natural patterns than the AFO. Unlike rigid AFOs, which restricted ankle motion and scored poorly for comfort and shoe compatibility similarly to previous studies ^18–20^, OrthoPed’s flexible design enhanced wearer satisfaction. Conversely, while Lycra socks allowed movement, they showed high variability in performance and were rated poorly for usability, particularly regarding ease of donning for younger children, reflective of previous studies ^21^.

OrthoPed combined the strengths of both comparators: a balance of support and mobility, strong functional performance, and the highest overall user satisfaction. Parents and children consistently preferred OrthoPed for its comfort, aesthetics, and ease of use, factors known to directly influence paediatric orthotic adherence ^22^. Notably, all parents perceived improvements in their child’s walking.

The immediate availability and low cost of OrthoPed present a significant opportunity to reduce health service expenditure and clinical workload by eliminating bespoke moulding and multiple fitting appointments. These features make it particularly promising for lower-resource settings or periods of high clinical demand.

Given these results, a multi-centre trial is now required to evaluate long-term clinical efficacy and cost-effectiveness. Such a trial should include longitudinal follow-up and health economic evaluations to determine if these preliminary functional gains translate to sustained system-level outcomes.

## Limitations

As a feasibility trial, this study was not powered to detect definitive statistical differences. The single-site design and small, heterogeneous sample limited subgroup analysis. Additionally, the protocol was demanding for younger participants, and short-term adaptation may have influenced comparisons. Future research should simplify protocols and incorporate longitudinal follow-up to evaluate sustained use in daily life.

## Conclusion

This study provides evidence that the OrthoPed splint is a safe, feasible, and highly acceptable intervention for paediatric foot drop. While recruitment was challenged by travel burdens, high compliance and positive user feedback support the viability of larger trials within standard care pathways. As a market-ready, adaptable solution, OrthoPed has the potential to reshape neurorehabilitation device provision globally.

## Ethical Approval

Ethical approval for this study was provided by Yorkshire & The Humber - Sheffield Research Ethics Committee, REC number: 23/YH/0004. The study was approved by the Health Research Authority along with confirmation of no objection from the Medicines and Healthcare Products Regulatory Agency. Parental consent and participant assent was provided for each participant prior to data being collected.

## Data sharing statement

Anonymised participant gait data are available on request by contacting the corresponding author.

## Data Availability

All data produced in the present study are available upon reasonable request to the authors

## REFERENCES

1. Miller Renfrew L, Lord AC, Warren J, Hunter R. Evaluating the Effect of Functional Electrical Stimulation Used for Foot Drop on Aspects of Health-Related Quality of Life in People with Multiple Sclerosis. Int J MS Care. 2019;21(4):173–182. doi:10.7224/1537-2073.2018-015

2. Meilahn JR. Tolerability and Effectiveness of a Neuroprosthesis for the Treatment of Footdrop in Pediatric Patients With Hemiparetic Cerebral Palsy. PM and R. 2013;5(6):503–509. doi:10.1016/j.pmrj.2012.11.005

3. Romkes J, Hell AK, Brunner R. Changes in muscle activity in children with hemiplegic cerebral palsy while walking with and without ankle-foot orthoses. Gait Posture. 2006;24(4):467–474. doi:10.1016/j.gaitpost.2005.12.001

4. Geboers JF, Drost MR, Spaans F, Kuipers H, Seelen HA. Immediate and long-term effects of ankle-foot orthosis on muscle activity during walking: A randomized study of patients with unilateral foot drop. Arch Phys Med Rehabil. 2002;83(2):240–245. doi:10.1053/apmr.2002.27462

5. Lam WK, Leong JCY, Li YH, Hu Y, Lu WW. Biomechanical and electromyographic evaluation of ankle foot orthosis and dynamic ankle foot orthosis in spastic cerebral palsy. Gait Posture. 2005;22(3):189–197. doi:10.1016/j.gaitpost.2004.09.011

6. Delafontaine A, Gagey O, Colnaghi S, Do MC, Honeine JL. Rigid Ankle Foot Orthosis Deteriorates Mediolateral Balance Control and Vertical Braking during Gait Initiation. Front Hum Neurosci. 2017;11. doi:10.3389/fnhum.2017.00214

7. Firouzeh P, Sonnenberg LK, Morris C, Pritchard-Wiart L. Ankle foot orthoses for young children with cerebral palsy: a scoping review. Disabil Rehabil. 2021;43(5):726–738. doi:10.1080/09638288.2019.1631394

8. Vinci P, Gargiulo P. Poor compliance with ankle-foot-orthoses in Charcot-Marie-Tooth disease. Eur J Phys Rehabil Med. 2008;44(1):27–31. http://www.ncbi.nlm.nih.gov/pubmed/18385625

9. Kirker S. Orthoses for neurological ankles. Pract Neurol. 2022;22(4):311–316. doi:10.1136/practneurol-2022-003357

10. Prenton S, Kenney LPJ, Cooper G, Major MJ. A sock for foot-drop: A preliminary study on two chronic stroke patients. Prosthet Orthot Int. 2014;38(5):425–430. doi:10.1177/0309364613505107

11. Chambers TL. Seven questions about paediatric research. J R Soc Med. 2000;93(6):320–321. doi:10.1177/014107680009300615

12. Enright PL. The Six-Minute Walk Test Introduction Standards and Indications 6-Minute Walk Test Versus Shuttle Walk Test Safety Variables Measured Conducting the Test Ensuring Quality Factors That Influence 6-Minute Walk Distance Interpreting the Results Improving the 6-Minute Walk Distance Summary. 2003.

13. Springer BA, Marin R, Cyhan T, Springer BA. Normative Values for the Unipedal Stance Test with Eyes Open and Closed. Journal of Geriatric Physical Therapy. 2007;30:7.

14. Williams EN, Carroll SG, Reddihough DS, Phillips BA, Galea MP. Investigation of the timed ‘Up & Go’ test in children. Dev Med Child Neurol. 2007;47(8):518–524. doi:10.1111/j.1469-8749.2005.tb01185.x

15. Daly JJ, McCabe JP, Gor-García-Fogeda MD, Nethery JC. Update on an Observational, Clinically Useful Gait Coordination Measure: The Gait Assessment and Intervention Tool (G.A.I.T.). Brain Sci. 2022;12(8):1104. doi:10.3390/brainsci12081104

16. Varni JW, Burwinkle TM, Seid M. The PedsQL^TM^ as a pediatric patient-reported outcome: reliability and validity of the PedsQL^TM^ Measurement Model in 25,000 children. Expert Rev Pharmacoecon Outcomes Res. 2005;5(6):705–719. doi:10.1586/14737167.5.6.705

17. Avery KNL, Williamson PR, Gamble C, et al. Informing efficient randomised controlled trials: exploration of challenges in developing progression criteria for internal pilot studies. BMJ Open. 2017;7(2):e013537. doi:10.1136/bmjopen-2016-013537

18. van der Wilk D, Hijmans JM, Postema K, Verkerke GJ. A user-centered qualitative study on experiences with ankle-foot orthoses and suggestions for improved design. Prosthet Orthot Int. 2018;42(2):121–128. doi:10.1177/0309364616683981

19. Silva PC, Silva MT, Martins JM. Evaluation of the contact forces developed in the lower limb/orthosis interface for comfort design. Multibody Syst Dyn. 2010;24(3):367–388. doi:10.1007/s11044-010-9219-6

20. Mulroy SJ, Eberly VJ, Gronely JK, Weiss W, Newsam CJ. Effect of AFO design on walking after stroke: Impact of ankle plantar flexion contracture. Prosthet Orthot Int. 2010;34(3):277–292. doi:10.3109/03093646.2010.501512

21. Rennie DJ, Attfield SF, Morton RE, Polak FJ, Nicholson J. An Evaluation of Lycra Garments in the Lower Limb Using 3-D Gait Analysis and Functional Assessment (PEDI). Vol 12. 2000. www.elsevier.com/locate/gaitpost

22. Bashir AZ, Dinkel DM, Pipinos II, Johanning JM, Myers SA. Patient Compliance With Wearing Lower Limb Assistive Devices: A Scoping Review. J Manipulative Physiol Ther. Elsevier Inc. 2022;45(2):114–126. doi:10.1016/j.jmpt.2022.04.003

